# A genome-wide association analysis of loss of ambulation in dystrophinopathy patients suggests multiple candidate modifiers of disease severity

**DOI:** 10.1101/2021.11.03.21265887

**Authors:** Kevin M. Flanigan, Megan A. Waldrop, Paul T. Martin, Roxane Alles, Diane M. Dunn, Lindsay N. Alfano, Tabatha R. Simmons, Melissa Moore-Clingenpeel, John Burian, Sang-Cheol Seok, Robert B. Weiss, Veronica J. Vieland

## Abstract

The major determinant of disease severity in patients with severe Duchenne muscular dystrophy (DMD) or milder Becker muscular dystrophy (BMD) is whether their dystrophin gene (*DMD*) mutation disrupts the mRNA reading frame or allows expression of a partially functional protein. However, even in the complete absence of dystrophin, variability in disease severity is observed, and candidate gene studies have implicated several genes as possible modifiers. Our previous genome-wide association study (GWAS) for age at loss of ambulation (LOA) in DMD provided confirmation for the role of genetic modifiers of TGF-β signaling in disease progression. Here we present the largest genome-wide search to date for loci influencing disease severity in DMD patients. Availability of subjects for such studies is still quite limited, leading to modest sample sizes, which present a challenge for GWAS design. We have therefore taken special steps to minimize heterogeneity within our dataset at the *DMD* locus itself, taking a novel and conservative approach to mutation classification to effectively exclude the possibility of residual dystrophin expression. We have also utilized statistical methods that are well adapted to smaller sample sizes, including the use of a novel linear regression-like residual for time to ambulatory loss and the application of evidential statistics for the GWAS approach. Based on the resulting sample size of N = 419 patients, we have identified multiple potential candidate genetic modifier loci. In a companion paper to this one, we use a systematic bioinformatic pipeline to implicate specific genes within these loci as potential DMD modifiers.

## Introduction

The X-linked dystrophinopathies result from mutations in *DMD*, the largest known human gene spanning ∼2.2 Mb of the X chromosome with 79 exons encoding the 427 kDa muscle isoform of the dystrophin protein. Dystrophin links the cytoskeleton with the sarcolemmal dystrophin-associated glycoprotein (DAG) complex, via N-terminal and central rod region actin binding domains, and a C-terminal sarcoglycan binding domain. The DGC complex is in turn linked to ligands in the extracellular matrix, allowing stabilization in the presence of the forces associated with muscle contraction. The absence of dystrophin results in membrane fragility, leading to the myofiber degeneration, inflammatory responses, tissue fibrosis and fatty replacement that are the histopathologic correlates of Duchenne muscular dystrophy (DMD). This typically leads to loss of ambulation (LOA) by ∼12 years and to death due to cardiac or pulmonary insufficiency by the third decade of life. In contrast, Becker muscular dystrophy (BMD) is associated with expression of a partially functional protein, which most often is the product of a mutation that results in a frame-preserving deletion of one or more exons within the central rod domain of dystrophin and preservation of functional N- and C-terminal protein binding domains. Genomic mutation analysis is often accompanied by interpretations of the predicted reading frame, which is used by clinicians to predict phenotype. Exceptions to this “reading frame rule” as interpreted from genomic DNA frequently occur, with most of them explained by molecular mechanisms that restore an open reading frame in the mRNA ^1^.

Although functional dystrophin level is a primary determinant of disease severity in the dystrophinopathies, there is still variation in severity among DMD patients who lack the protein altogether. Historically, age at LOA in DMD patients ranged from around age 7 to age 12 years, and although the use of corticosteroids has changed the natural history slightly, adding 1-3 years to ambulation, a significant range remains over which ambulation may be lost. Environmental factors may influence this range, but genetic modifiers are also likely to play a role. Identifying modifier genes in patient cohorts has relied on the successful utilization of animal models of muscular dystrophy, including the *mdx* mouse model which carries a *Dmd* mutation, as well as models that disrupt the DAG complex. Numerous studies have demonstrated that proteins involved in inflammation, fibrosis, regeneration, and sarcolemma repair modify the dystrophic process in these animal models. Linkage studies in the DBA/2J genetic background identified a protein polymorphism in latent TGF-β binding protein 4 (*Ltbp4*) associated with increased levels of TGF β -mediated fibrosis ^2^. Gene expression studies identified the matricellular protein osteopontin (*Spp1*) as highly expressed in dystrophic skeletal muscle in mouse, dog, and humans, where it contributes to the balance between inflammatory, regenerative, and fibrotic processes ^3^. Based on this animal model work, candidate gene association studies in human DMD patients have shown that common regulatory SNPs in the proximal promoter of *SPP1* ^4^ and a common *LTBP4* haplotype with four non-synonymous SNPs in strong linkage disequilibrium are associated with disease severity ^5^. In these studies, support for *SPP1* and *LTBP4* SNPs are stronger with dominant or recessive models, and cohort effects are evident with variable success at replication ^6^. Follow-up studies have confirmed the predicted effect of the “protective” and “risk” human LTBP4 polymorphisms on the dystrophic phenotype in mice and have shown epistatic interactions between *Spp1* and *Ltbp4* in modulating fibrosis through pathological TGF β signaling ^7^. The role of the TGF β pathway in modifying dystrophic pathology in mice is also supported by the identification of annexin A6 (*Anxa6*), a Ca2+-binding protein involved in sarcolemma repair, as a downstream effector of TGF β signaling ^7^, and with alleles of *Ltbp4* and *Anxa6* acting jointly to modify the dystrophic phenotype. Extreme phenotype study designs have also been used. Long-term ‘escapers’ from early loss of ambulation in the golden retriever muscular dystrophy (GRMD) dog model suggested *Jagged1* as a modifier gene, supporting earlier work that the Notch signaling pathway is perturbed in *mdx* satellite cell self-renewal and quiescence ^8^.

As an extension of candidate modifier gene studies in DMD patients, there have been two previous reports of GWAS for age at loss of ambulation. The first was based on an exome chip (27K markers) and 109 ancestrally homogeneous samples ^9^. While no loci reached statistical significance, subsequent filtering for genes in candidate pathways identified a haplotype that included a known functional SNP, rs1883832, in the Kozak sequence of the *CD40* ^4,9^ gene, which encodes a costimulatory receptor mediating immune and inflammatory responses in a variety of cell types. More recently, we reported a preliminary GWAS using 253 non-ambulant subjects ^10^ and observed two loci above the genome-wide significance threshold using the recessive model suggested by the prior candidate gene study. One locus was *LTBP4* itself, where fine mapping revealed *cis*-eQTL variants in linkage disequilibrium with “protective” IAAM LTBP4 protein isoform and associated with *LTBP4* mRNA expression. The other locus was a distal enhancer containing *cis*-eQTL variants associated with thrombospondin-1 (*THBS1*) expression, a matricellular protein with multifunctional properties including promoting TGFβ signaling through latent protein complex activation ^11^. Finally, a recent two-stage design using whole exome sequencing of extreme phenotypes followed by replication identified *TCTEX1D1* as an LOA modifier ^12^. This study highlighted the importance of novel study designs in searching for modifiers of DMD, but also emphasized the need for more complete knowledge of mutation-specific effects that may increase residual dystrophin levels.

The current work is distinct from our previous GWAS study in several critical ways. First, we have substantially increased the cohort to N = 419 dystrophinopathy patients, and while previously we analyzed only those subjects who had lost ambulation, here we utilize survival analysis to include subjects who are still ambulatory at last assessment. Second, we have used a more conservative approach to excluding subjects whose *DMD* mutations may produce residual dystrophin, to minimize confounding of modifier gene effects with LOA variation due to mutations within the *DMD* gene itself. Third, instead of the usual (frequentist) approach to assessing evidence for genetic association, we have utilized statistical methods that are well adapted to analysis in smaller data sets, as we describe below. This is critical for the study of a rare disease like DMD, for which the accrual of data sets on the order of typical GWAS designs (thousands or even tens of thousands of subjects) is not feasible.

Our current analysis implicates several novel regulatory loci as potential modifiers of DMD severity. In the companion paper [citation] we utilize a systematic *in silico* pipeline integrating multiple data sources to evaluate the biological plausibility of these candidate modifiers as a first step toward further studies to determine pathogenesis.

## Materials and Methods

### Clinical Classification

All subjects have a known *DMD* mutation confirmed by either MLPA or DNA sequencing, as has been previously described ^13,14^. The study was conducted under research protocols approved by the Nationwide Children’s Hospital Institutional Review Board (Approval #0502HSE046) and the University of Utah Institutional Review Board (Approval #IRB_00094017). Written informed consent was obtained from the parent/guardian of each minor subject, or from the subject themselves if 18 years of age or older. Genomic DNA was extracted from venous blood samples and phenotypic data were extracted from clinical records; most subjects underwent prospective clinical examinations as well.

Ambulatory loss was defined by full-time wheelchair use (requiring wheelchair within the home) and LOA was recorded to the nearest month when available, and otherwise to the nearest half-year of age. Glucocorticoid treatment (Steroid Y) was defined as use of any steroid regimen for more than six months that began at least six months prior to loss of ambulation. Untreated (Steroid N) was defined as never exposed, exposed to any steroid regimen for less than six months, or onset of treatment less than six months prior to loss of ambulation. All genotyped subjects had complete clinical data, including information on steroid use.

### Subjects in the final cohort

The study design is shown in Fig 1A-D. In previous studies, we used physician-assigned diagnoses of DMD and BMD, which are themselves defined clinically by age at LOA, and therefore excluded individuals who still ambulated at age 20. Here we include patients regardless of their age at LOA but in order to minimize the impact of trace dystrophin protein expression ^15^, we classified *DMD* mutations using a strict loss-of-function (LoF) definition from predicted muscle-isoform transcription, splicing, and translation of their *DMD* mutation (Fig 1A). We excluded typical Becker mutations, for example, “in-frame” deletions in the central rod domain, nonsense mutations in exon 1 that use a downstream translation initiation site ^16^, and “out-of-frame” exon 3 to 7 deletions ^17^. However, we also excluded patients with mutations that typically have been reported as “out-of-frame” or nonsense mutations on clinical reports but may occasionally result in low-level dystrophin expression based upon a known or suspected mechanism for bypassing frame-truncating *DMD* mutations. Excluded mutations from this partial LoF class include “out-of-frame” exon 45 deletions, which sometimes can provoke low-level exon 44 skipping that restores the reading frame, as well as exon 8 “skippable” mutations ^18^. We and others have previously demonstrated that exons 23-42 constitute a “zero-frame context,” in which deletion of any exon(s) results in an open reading frame. Nonsense mutations within these exons—although classically categorized as frame-truncating—can be associated with exon skipping due to poorly-predicted damage to exon definition elements recognized by the spliceosome that restores the open reading frame by excluding the mutated exon from the mature mRNA^1,19,20^. Because of this strict LoF mutation criteria, we included 311 non-ambulant LoF subjects, but excluded 166 non-ambulant partial LoF subjects, of which 132 had lost ambulation by 20 years (Fig 1A).

**Figure 1.**
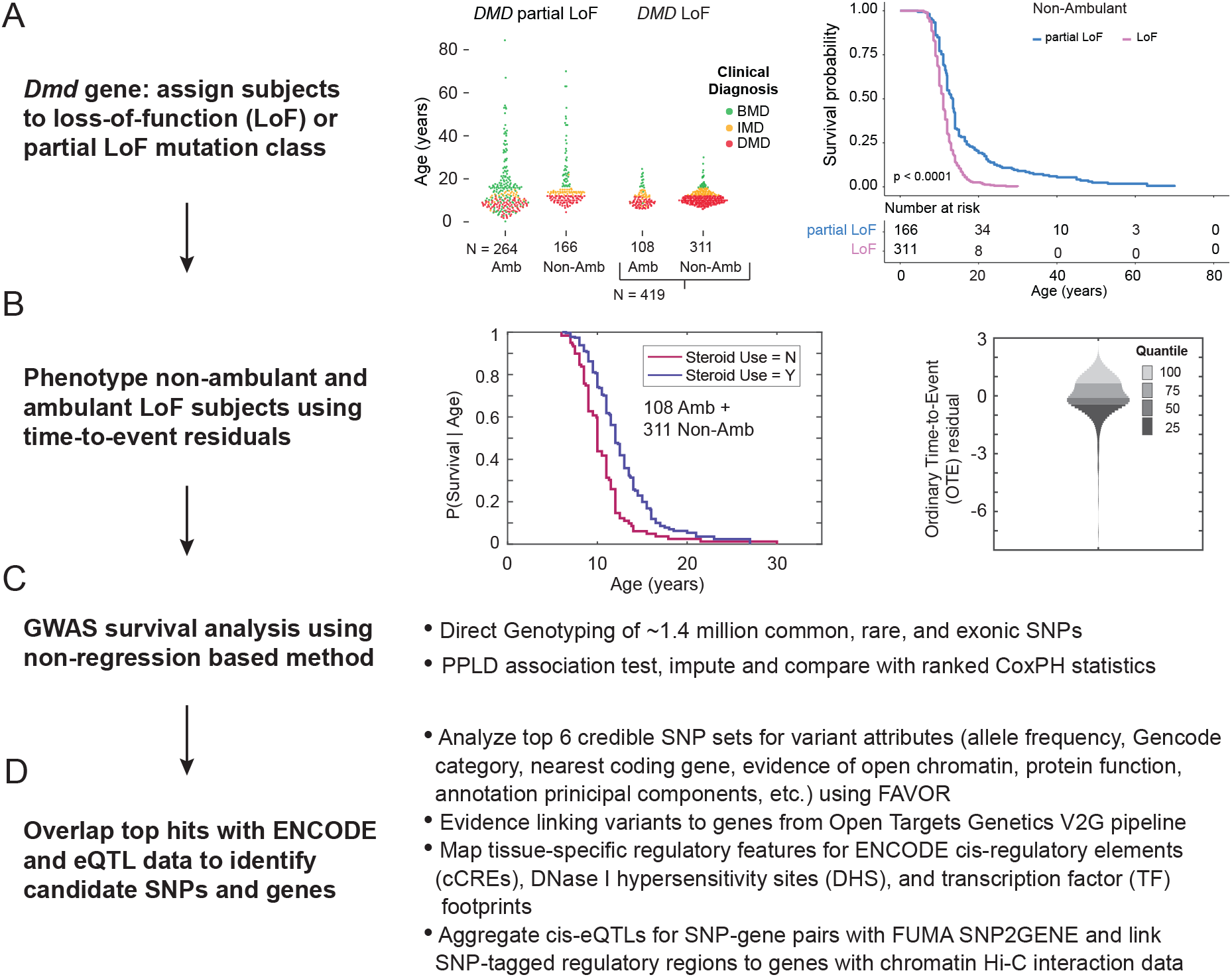
Outline of the analyses performed in this study. **(A)** DMD mutation class and LOA cohort selection. The middle panel shows the UDP cohort divided by dystrophin loss-of-function class with the y-axis is age at loss of ambulation for non-ambulant (Non-Amb) patients or age at last visit for ambulant (Amb) patients. Individual patients are colored by their clinical diagnosis: DMD (red), IMD (orange), and BMD (green). The right panel shows LOA survival curves for non-ambulant patients. **(B)** The middle panel shows the Kaplan-Meier curves in the censored LoF cohort (N = 419 patients)> The right panel shows a density plot of the OTE residuals for the study cohort with the unit on the y-axis in deviations in age at LOA from expectation, after adjusting for steroid use, on the standard normal scale. Note that residuals are oriented such that smaller (negative) values represent older-than-expected LOA, while larger (positive) values represent younger-than-expected LOA. **(C)** GWAS study design and **(D)** bioinformatic pipeline for annotating and linking variants to genes.

After this filtering step, the final cohort comprised 201 distinct *DMD* mutations, encompassing 71 multi-exon deletions, 52 nonsense, 23 multi-exon duplications, 21 frameshifts, 14 single exon deletions, 11 single exon duplications, 6 splice donor, and 3 splice acceptor mutations (see S1 and S2 Tables). Recent observations of patients with exon 2 duplications catalogued within the UDP database suggests that many of these patients may have delay in LOA in relation to other mutations, perhaps due to utilization of a downstream internal ribosome entry site to initiate translation of a highly functional dystrophin; as these results have not yet been published or validated, Dup2 patients were not excluded from the LOF group, but are discussed below.

In the process of genotype cleaning we dropped 2 individuals for an excess (> 4%) of missing genotypes; 4 individuals from 4 MZ twin pairs; and 43 individuals still ambulant with last assessment at ≤ 6 years of age (the minimum observed LOA in our cohort) and not removed for other reasons. Relationships among individuals were determined using KING (Wei-Min Chen, http://people.virginia.edu/∼wc9c/KING) and confirmed against clinical records; in addition to the MZ twins previously mentioned, the data set included 9 full-sibling pairs, 5 half-sibling pairs, 1 set of 3 half-sibs, 1 first cousin pair and 1 set of 3 first cousins. The SmartPCA routine from EIGENSOFT 6.0.1^32^ was used to determine ancestral outliers among the unrelated individuals, using the default selection criteria (10 principal components, 6 standard deviations for flagging outliers along those components); this suggested that 50 of the remaining individuals were likely to be outliers. These individuals were removed prior to analyses. This left 419 individuals for analysis. The proportion of subjects still ambulatory (censored) as of last examination was 26.7%. 69.8% of subjects had been treated with steroids, and 30.2% were untreated.

For additional exploration of ancestry, we used the filtered Illumina 650Y array genotype data set of 938 unrelated individuals from the CEPH-Human Genome Diversity Panel (HGDP) as reference populations ^21^. SNPs in common with the Illumina Omni2.5Exome genotype data from UDP subjects were merged and filtered to remove SNPs in high LD using the plink “--indep-pairwise 50 10 0.1” parameter. The LASER/TRACE 2 server (https://laser.sph.umich.edu) was used to determine principal components from the genotype data, and for each study subject, we identified the 10 nearest reference individuals from 1,385 individuals in the Europe imputed POPRES 3 reference panel based on Euclidean distance using the first 10 principal components. For ancestry exclusion, we used the Z score threshold > 6, proposed by Taliun et al. (2017) ^22^ to identify outliers among study sample by estimating the similarity of each study individual to the 10 nearest neighbors in the European POPRES reference. For visualization, population structure was estimated using the ADMIXTURE software ^23^ with the clustering parameter K = 6. The output of multiple ADMIXTURE runs (n=10) was summarized and plotted using the post-processing tool, pong ^24^. The composition of the HGDP 938 and DMD study populations assuming K = 6 source populations is shown in S1A Figure. The 419 DMD subjects were divided into two groups: n = 389 and n = 30 using a Z score threshold of 6 for estimating the similarity to POPRES European reference samples (S1B Fig).

### Illumina Array Genotyping and quality control

Genomic DNAs were genotyped on Illumina Infinium Omni2.5Exome-8 v1.3 and v1.4 BeadChip arrays as previously described ^10^. Of the initial set of 2,562,265 SNPs, 337,521 monomorphic sites were removed prior to analysis, as were 1,837 for low genotyping rate (< 95%) and 46,306 duplicate markers. We also checked for excess missingness (≥ 10%) and for excess heterozygosity (≥ 10%) in the non-pseudoautosomal region of X, but no SNPs were dropped on the bases of these checks; any remaining heterozygous genotypes on X were set to missing, and 464 Y and mitochondrial SNPs were also removed. Due to the small sample sizes, we imposed a MAF threshold ≥ 3% (which removed 791,787 SNPs), and a HWE threshold of P < 1E-05 (which removed 777 SNPs). This left a total of 1,383,573 SNPs in the analysis. As an additional quality control step, 322 samples with *DMD* single or multiexon deletion / duplication mutations had their breakpoints verified from the log_2_ normalized probe intensity R ratio within the *DMD* locus.

### Genotype Imputation and Haplotype Phasing

PLINK binary files were converted to VCF format using the Haplotype Reference Consortium release 1.1 (HRC.r1-1) sites and the HRC preparation checking tool (https://www.well.ox.ac.uk/∼wrayner/strand/) to check strand, alleles, position, and REF/ALT assignment. Genotype imputation and haplotype phasing utilized the Sanger Imputation Service (https://imputation.sanger.ac.uk) running EAGLE2 pre-phasing and the PBWT imputation pipeline with the HRC (r1.1) reference panel. Imputed SNPs were quality filtered using bcftools, only keeping SNPs with an IMPUTE2 info score parameter ≥ 0.8.

### Statistical Methods

The primary GWAS analysis was based on the posterior probability of (trait-marker) linkage disequilibrium (PPLD), computed using the software package KELVIN ^25^ to assess evidence for or against association between LOA and each SNP in turn. The PPLD was selected for these analyses for several reasons: (i) it allows us to retain related individuals in the data set along with unrelateds; (ii) it can detect genotypic effects on trait variances as well as on means; (iii) it is essentially “model-free,” making minimal distributional assumptions (as described below) and bypassing the need for separate analyses under recessive, additive and dominant models, which can have highly inflationary effects in conventional regression-based approaches ^26^; and (iv) it is designed to measure the strength of statistical evidence, for each SNP in turn, either for an association with LOA or against an association with LOA. This last feature may be critically important in the context of smaller sample sizes, where distinguishing weak evidence in favor of association from evidence against association boosts the separation of “signal” from “noise.” The advantages of the PPLD over regression analysis in the context of GWAS based on small to moderate sample sizes are detailed in ^27^.

In order to adjust for steroid treatment, a known modifier of LOA, we pre-processed the phenotypes using conventional survival analysis including steroid treatment as a covariate, extracting Ordinary Time-to-Event (OTE) residuals, so called because they can be interpreted in the same way as the residuals one obtains from “ordinary” linear regression ^28^ —that is, as estimates of how far from expectation is an individual’s LOA (whether observed or censored), given the individual’s steroid treatment status. These residuals were then used as the phenotype for PPLD analysis. Residuals were based on fitting a 2-parameter Weibull distribution, separately in the two steroid groups. The Weibull distribution was fit based on unrelated individuals in each group; the resulting regression equation was then used to assign OTE values to relatives.

Our approach allows us to mitigate a major limitation of standard regression-based approaches to GWAS in moderately sized samples. As is well known, regression methods, which focus on group mean effects, are dependent on adequate numbers of individuals per group. A standard LOA analysis would need to include both steroid use (generally dichotomized) and mutation class (sometimes dichotomized and sometimes broken down further by sets of specific mutations), along with genotype. By restricting our cohort to a strictly defined set of truncating mutations, and utilizing a non-regression-based method for the GWAS itself, this leaves us subdividing the dataset only by steroid when making the covariate adjustments (see below), and our current sample size is more than adequate to allow good estimation of the survival curve – and therefore the residuals – separately in each steroid group.

The PPLD is computed as follows. At each SNP, the PPLD is parameterized as a mixture of 3 normal LOA distributions, 1 per SNP genotype (11, 12, 22), and model-averaging over the 6 trait parameter parameters (*μ*_11_, *μ*_12_, *μ*_22_, *σ*_11_, *σ*_12_, *σ*_22_) was used to calculate a Bayes ratio (BR), or integrated likelihood ratio under an essentially uniform prior ^29^, assuming Hardy-Weinberg equilibrium. This parameterization makes only the weak assumption that remaining variation in LOA after accounting for genotype (and steroid status) is approximately normally distributed, as we have shown elsewhere ^29,30^ that the PPLD is highly robust to even large departures from normality. The trait allele frequency was set equal to the SNP MAF to align with standard GWAS analyses, which assess the potential impact of SNP genotype on the phenotype directly, rather than modeling this relationship explicitly via an underlying (unmeasured) trait locus in LD with the SNP. Related individuals were handled in the association analyses via KELVIN’s built-in Elston-Stewart algorithm for likelihood calculations on pedigrees.

Letting H_A_ be the hypothesis “phenotypes depend on SNP genotype,” and H_0_ be the hypothesis “phenotypes are independent of SNP genotypes,” the BR expresses how much more probable are the data on H_A_ compared to H_0_. For convenience, we then convert the BR to the posterior probability scale (0,..,1) using Bayes’ theorem. Here we use a prior probability of *π* = 0.0004, based in part on an empirical estimate of the prior probability that any two genomic positions would be in LD. ^30^ Thus PPLD > 0.0004 indicates (some degree of) evidence for association, while PPLD < 0.0004 indicates evidence against association. Note that whatever choice is made for *π*, when the BR > 1 the PPLD > *π*, indicating (some degree of) evidence in favor of association, while when the BR < 1 the PPLD will be < *π*, indicating (some degree of) evidence against association. Thus *π* operates as a scaling factor, but it does not fundamentally affect the interpretation of the PPLD.

The PPLD is an evidence measure, designed in keeping with the “evidentialist” school of statistical inference. Because evidentialism is less familiar to many in the genetics community, and to aid in interpretation of the results, here we comment briefly on key features of evidentialist evidence measures; for fuller discussion of the evidentialist program in statistics see ^31-36^. In frequentist settings, a significance threshold is set in advance for the p-value, and only SNPs with p-values surpassing that threshold are considered significant. The p-value = P(D | H_0_), where D represents the data, and setting a low significance threshold is an attempt to protect against the possibility of declaring significance when the posterior probability P(H_A_ | D) is low. However, as is well known, 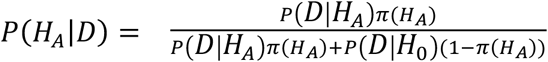, which is a function of not only P(D | H_0_) but also P(D | H_A_) and the prior probability *π*(H_A_) of association. Thus, even a small p-value can correspond to a low posterior probability of association in some circumstances. The PPLD, by contrast, is a direct estimate of the posterior probability of association, taking into account H_0_, H_A_ and *π*(H_0_). Hence, there is no need to compare it to a significance threshold to guard against false positive findings. For closely related reasons, the PPLD is not subject to multiple testing “corrections” as are p-values. The need to correct the p-value arises because it is used to represent both an error probability and the strength of evidence. By contrast, the BR is a classical evidentialist or Bayesian evidence measure, which decouples the estimate of the strength of evidence from the error probabilities associated with decisions regarding significance ^31-34,36^ ; it is not itself an error probability and is therefore not subject to multiple testing correction. Therefore, in following up on findings we start with the highest PPLDs and work our way down the list. The PPLD is then treated as one key piece of information in assessing the overall likelihood that a SNP is in fact modifying the DMD phenotype (or in LD with a modifier locus), taken together with consideration of ancillary information, as described below.

### Fine Mapping

We stratified directly genotyped SNPs by their PPLD score and selected those with PPLD ≥ 0.4 for detailed analysis. This threshold is somewhat arbitrary, but it effectively separated a very small number of SNPs for detailed follow up. Regional PPLD values for imputed SNPs ±750 kb from the lead SNP were calculated and regional LocusZoom association plots were generated using the locuszoom.org tool. We defined the credible SNP set in each region as markers with a PPLD ≥ 0.05 ±750 kb from the lead SNP, loosely following methods developed for use with GWAS p-values ^37^.

## Results

Kaplan-Meier estimates of the survival function S (time to LOA) based on the N = 419 LoF cohort are shown in Fig 1B, separately for the two steroid treatment groups. The estimated mean (standard deviation) of LOA is 10.1 (2.6) and 12.3 (3.4) for the steroid N and Y groups, respectively. As can be seen, there is considerable variability in LOA, with no individuals losing ambulation prior to age 6, but a substantial number (44) still walking beyond age 15. The distribution of the Ordinary Time-to-Event (OTE) residuals as the phenotypic values used as input for PPLD analysis are also shown in Figure 1B.

### GWAS for loss of ambulation (LOA)

Fig 2A shows the primary genome-wide scan results of the PPLD analysis. 94.4% of SNPs returned a PPLD < 0.0004, indicating evidence against association with LOA. There were 74 (0.0053%) SNPs with PPLD ≥ 0.04, 28 (0.0020%) with PPLD ≥ 0.10, and 8 (0.0006%) with PPLD ≥ 0.40; this last group represented 6 distinct loci. Table 1 shows the list of 19 distinct loci with PPLD ≥ 0.10, indicating the peak genotyped SNP as well as the maximum PPLD value in each region including imputed genotypes (Table 1). While imputation increased the maximum PPLD at some loci, it did not change the set of loci with PPLD ≥ 0.40; it did however somewhat shuffle the rank-ordering within this set. For comparison, Table 1 also shows the p-value and rank for these top SNPs from a Cox proportional-hazards regression (CPH) using the OTE residuals and an additive model. Although this is an “apples and oranges” contrast (again, see ^27^ for a detailed comparison of PPLD to CPH analysis), it does indicate that 5 out of 6 loci with PPLD ≥ 0.40 have some level of support from CPH regression. Note that the relatively “sparse” appearance of the PPLD Manhattan plot (Fig 2A) is due in part to differences in proportionality of pairwise *r*^*2*^ LD with the lead SNP and the test statistics as shown in LocusZoom plots of the rs12657665 region on chr5 (Fig 2B); the PPLD also produces fewer false positive signals than conventional regression analysis ^27^. We also evaluated the robustness of these top PPLD values after removing 30 subjects with a stricter definition of ancestry (S1 Fig) or excluding 12 subjects with *DMD* Dup2 mutations, as discussed above. Large decreases in PPLD values were observed for three loci after further ancestry filtering and two loci after Dup2 filtering (Table 1). Four of the top six loci were unaffected by the additional filtering criteria, while two of the top six loci (chr8 rs72681143 and chr5 rs10077875) showed a moderate decrease in PPLD values after Dup2 exclusion; note that the resulting PPLDs were still considerably higher than the prior probability of 0.0004.

**Figure 2.**
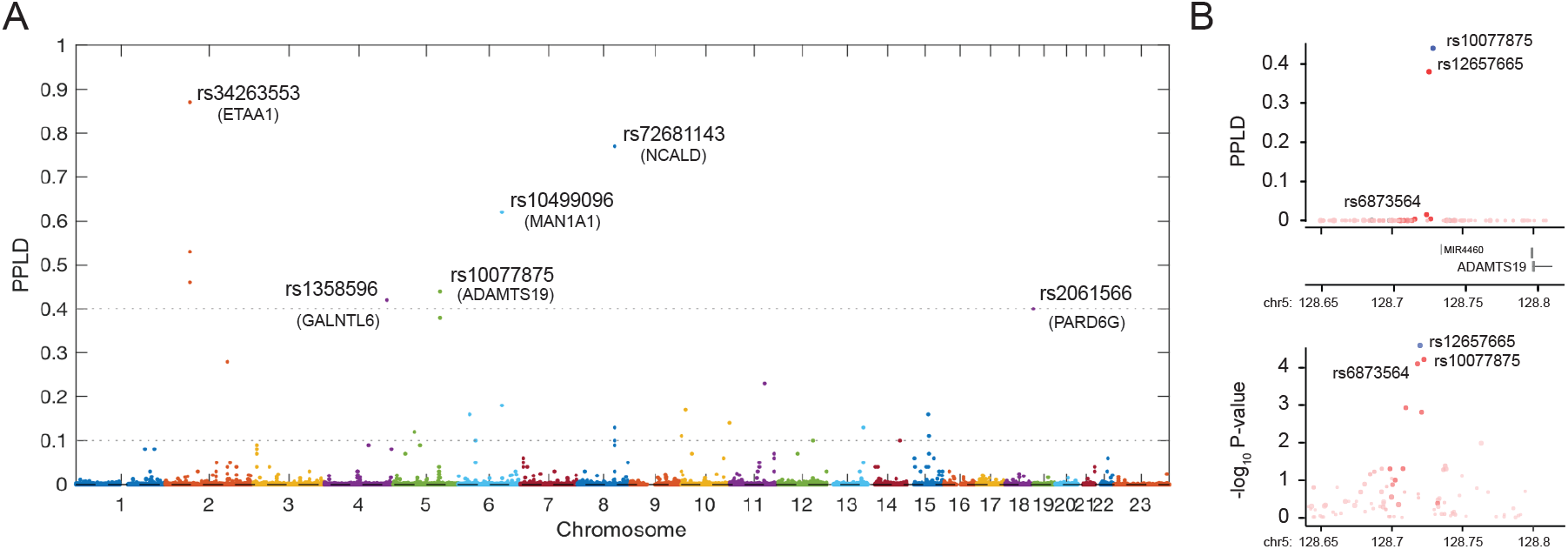
Manhattan plot of genome-wide scan. **(A)** The PPLD is on the probability scale and represents the estimated posterior probability that a SNP affects the phenotype, either directly or via linkage disequilibrium with one or more causal variants. Horizontal lines correspond to the heuristic cutoffs used to prioritize SNPs for further investigation in what follows. **(B)** LocusZoom plots of the lead SNP (rs10077875) from chr5 using PPLD values (upper panel) or p-values from a Cox Proportional Hazards regression with steroid exposure and SNP genotypes as covariates (lower panel).

**Table 1.** SNPs with PPLDs ≥ 0.10. Position: Physical positions are based on build 37. Marker: shown is the top genotyped SNP per region, where the regions are defined as in the main text. PPLD: The PPLD at the genotyped SNP. Max PPLD: The maximum PPLD in the region, which may occur at the originally genotyped SNP or at an imputed SNP. CPH Add_P: CPH Add_P: Cox Proportional Hazards Regression p-value with steroid exposure and SNP genotypes as covariates. CPH rank order: additive model ranked by p-value from 1,353,208 directly genotyped SNPs. Ancestry Filtered PPLD: 30 subjects with second criteria applied for ancestry filtering. Dup2 Filtered PPLD: 12 subjects removed with Dup2 *Dmd* mutations. V2G top gene: Variant to Gene nearest protein coding gene. Gencode: gencode annotation category for the Max marker.

| Chr | Position | Marker | PPLD | Max PPLD | CPH Add p-value | CPH rank order | Ancestry Filtered PPLD (n = 389) | Dup2 Filtered PPLD (n = 407) | V2G top gene | Gencode |
| --- | --- | --- | --- | --- | --- | --- | --- | --- | --- | --- |
| 2 | 67958890 | rs34263553 | 0.87 | 0.87 | 1.9E-03 | 4352 | 0.36 | 0.79 | ETAA1 | intergenic |
| 8 | 103189435 | rs72681143 | 0.77 | 0.83 | 0.45 | 642282 | 0.80 | 0.05 | NCALD | intergenic |
| 4 | 172432296 | rs1358596 | 0.42 | 0.75 | 1.2E-03 | 3016 | 0.65 | 0.54 | GALNTL6 | intergenic |
| 6 | 120491553 | rs10499096 | 0.62 | 0.64 | 3.0E-04 | 880 | 0.68 | 0.73 | MAN1A1 | intergenic |
| 5 | 128722696 | rs10077875 | 0.44 | 0.62 | 6.1E-05 | 239 | 0.89 | 0.03 | ADAMTS19 | intergenic |
| 18 | 77551738 | rs2061566 | 0.40 | 0.40 | 1.0E-05 | 50 | 0.50 | 0.22 | PQLC1 | intergenic |
| 2 | 171307693 | rs1816670 | 0.28 | 0.38 | 3.6E-04 | 1017 | 0.27 | 0.0075 | MYO3B | intronic |
| 11 | 97137756 | rs78973513 | 0.23 | 0.23 | 6.9E-06 | 31 | 0.25 | 0.09 | JRKL | intergenic |
| 10 | 13049885 | rs7921121 | 0.17 | 0.17 | 0.01 | 20370 | 0.13 | 0.11 | CCDC3 | intronic |
| 6 | 31205392 | rs3130530 | 0.16 | 0.14 | 0.10 | 158686 | 0.11 | 0.18 | HLA-C | intergenic |
| 15 | 59429111 | rs11539756 | 0.16 | 0.16 | 0.03 | 45865 | 0.15 | 0.0176 | MYO1E | UTR3 |
| 10 | 134349181 | rs11146413 | 0.14 | 0.08 | 3.7E-04 | 1048 | 0.0017 | 0.28 | INPP5A | intergenic |
| 13 | 101558219 | rs9513818 | 0.13 | 0.32 | 0.04 | 64644 | 0.23 | 0.0036 | TMTC4 | intergenic |
| 5 | 58272070 | rs62356653 | 0.12 | 0.12 | 1.3E-05 | 56 | 0.08 | 0.19 | PDE4D | intronic |
| 10 | 1449849 | rs4880848 | 0.11 | 0.11 | 0.90 | 1222924 | 0.0037 | 0.11 | ADARB2 | intronic |
| 15 | 61272080 | rs2140442 | 0.11 | 0.11 | 0.45 | 644035 | 0.10 | 0.2 | RORA | intronic |
| 6 | 47227944 | rs2281448 | 0.10 | 0.10 | 0.31 | 459744 | 0.0002 | 0.06 | TNFRSF21 | intronic |
| 12 | 94954104 | rs4761483 | 0.10 | 0.05 | 0.06 | 97480 | 0.11 | 0.0087 | PLXNC1 | intronic |
| 14 | 88421482 | rs113691242 | 0.10 | 0.07 | 6.5E-04 | 1646 | 0.11 | 0.0192 | GALC | intronic |

### Regional analysis of lead SNPs

Using a PPLD threshold ≥ 0.05 to define credible sets from the top 6 regions, 96 total SNPs were chosen for further analysis. Regional LocusZoom plots for these six loci are shown in Fig 3 and S2 Fig, with Kaplan-Meier survival estimates and genotype-specific OTE residual plots for each lead SNP. Note that survival curves that cross one another indicate evidence for genotypic effects on variances. Functional annotations of individual SNPs (S3 Table) revealed that all 96 SNPs were noncoding intergenic or intronic variants. Four of the credible sets displayed a very narrow range of pairwise SNP-to-SNP linkage disequilibrium (*r*^*2*^) of 0.96-0.98 for chr4 rs1358596 (Fig 3A), 0.90-0.96 for chr2 rs34263553 (Fig 3B), 0.89-0.98 for chr6 rs10499096 (S2A Fig), and 0.84-0.98 for chr5 rs12657665 (S2B Fig). The chr8 rs3899035 top SNP had a broader range of *r*^*2*^ values from 0.27-0.99 (S2C Fig), while the chr18 rs2061566 SNP was the only variant in the credible set as no other SNPs in the study sample had an *r*^*2*^ value > 0.3 (Fig 3C), similar to the rs2061566 LD pattern seen with European ancestry samples in the 1000 Genomes Project. Potential candidate genes within these regions are proposed in the companion paper.

**Figure 3.**
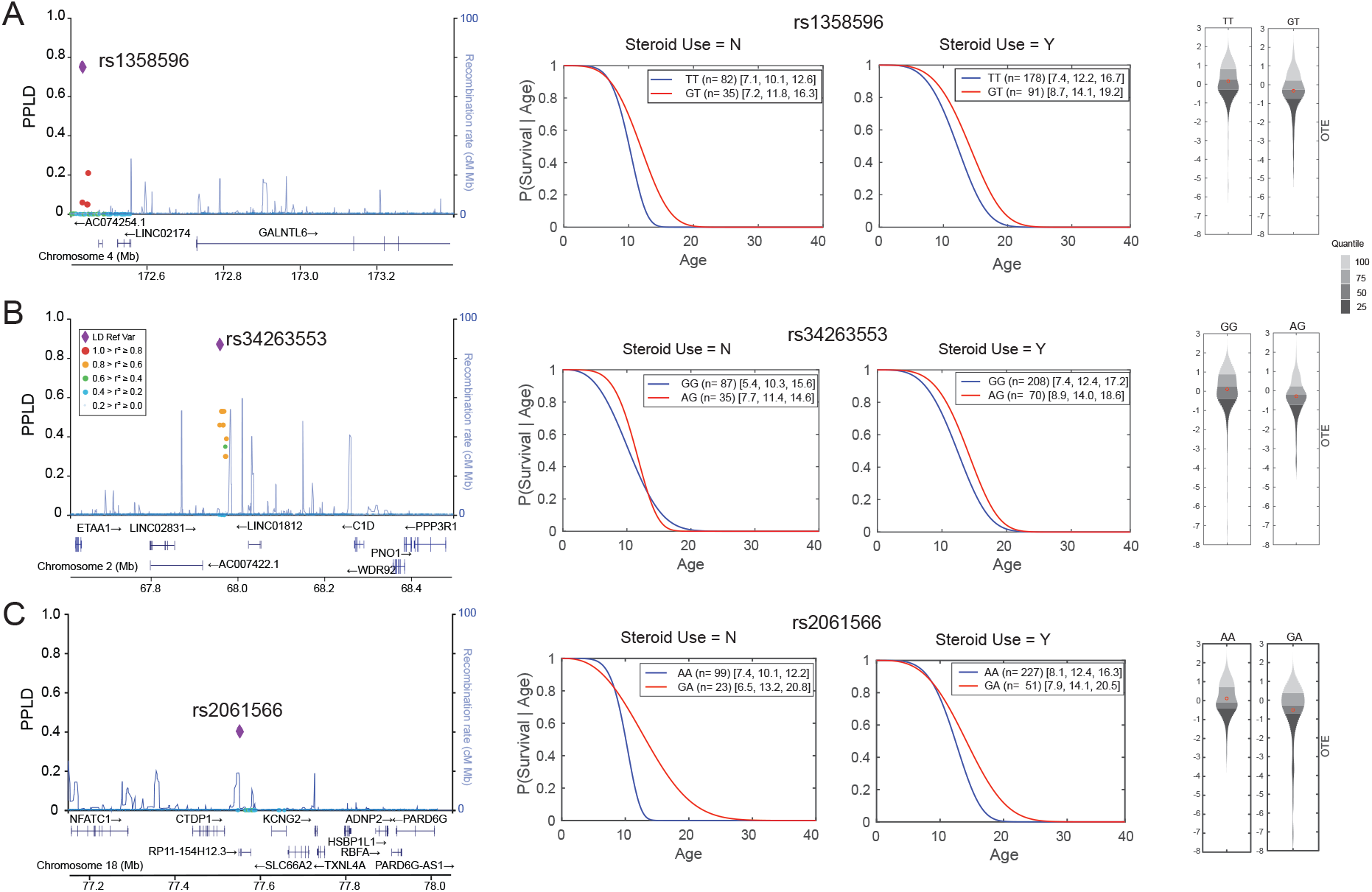
Select regional plots of loci with PPLDs ≥ 40%. **(A)** through **(C)** are three regions from Table 1 with PPLDs ≥ 40%. The **left panel** shows regional LocusZoom plots of SNPs with their PPLD values shown on the primary y-axis and colored by their LD (*r*^*2*^, red ≥ 0.8, orange ≥ 0.6, green ≥ 0.4, light blue ≥ 0.2) with the lead SNP indicated with a purple diamond. The **middle panel** shows the estimated (Weibull) survival curves for the two Steroid (N/Y) groups, respectively; legends include the number n of individuals followed by [10^th^ percentile, median, 90^th^ percentile] of the age at LOA distribution. The right panel shows the density plots for the OTE residuals with the y-axis units in deviations in age at LOA from expectation, after adjusting for steroid use. Note that residuals are oriented such that smaller (negative) values represent older-than-expected LOA, while larger (positive) values represent younger-than-expected LOA. Red dots indicate the mean within each group. Genotypes with <15 individuals are not shown, since our ability to accurately estimate the distribution in very small groups is limited.

## Discussion

In the largest genome-wide association study to date of the LOA phenotype in DMD subjects, we report the identification of multiple loci that may harbor genetic modifiers of DMD severity. Our approach was distinct from previous efforts, as it is based on a cohort carefully restricted—using highly conservative criteria—to truncating DMD mutations and uses statistical methods particularly well-adapted to GWAS in moderate sample sizes.

One aspect of these new SNP associations is that we did not detect previously identified modifier genes, which are reviewed in detail elsewhere ^6^. The PPLDs at these SNPs were all at or below the prior probability of SNP-trait association (Table 2). For comparison, we also show MOD scores, which are log_10_ maximum likelihood ratios (where the maximization is over the 6 parameters of the trait model); –log_10_(p-values) from CPH, and from variance-components regression (as we previously reported ^10^). The MODs range from 0.90 – 2.98. By contrast, MODs at the SNPs showing the PPLDs ≥ 0.40 (Table 1) range from 6.6 – 8.4. Table 2 clearly indicates that failure to detect these SNPs in the current analysis is not a consequence of our use of the PPLD; rather, evidence for these SNPs is simply not found in this particular cohort.

**Table 2.** Results at previously reported LOA-associated SNPs. *This SNP was imputed; the remainder were directly genotyped. There were no additional salient results at any of these loci in our dataset. ^1^Cox Proportional Hazards Regression with steroid exposure and SNP genotypes as covariates; shown here is the maximum –log_10_(p-value) over recessive (rec), additive (add), dominant (dom) models, with the maximizing model as indicated (relatives have been removed from the dataset). ^2^Variance-Components analysis run in Mendel under the Ped-GWAS option (allowing for inclusion of relatives) with steroid exposure and SNP genotypes as covariates, under the recessive model.

| Gene | SNP | PPLD | MOD | CPH<sup>1</sup> | Variance Components<br>QTL Analysis<br>(recessive)<sup>2</sup> |
| --- | --- | --- | --- | --- | --- |
| | | | | $-\log_{10}(\text{p-value})$ | $-\log_{10}(\text{p-value})$ |
| **SPP1** | rs28357094* | 0.0003 | 2.98 | 2.42 (dom) | 0.19 |
| **LTBP4** | rs710160 | 0.0002 | 0.97 | 1.16 (rec) | 0.17 |
| **LTBP4** | rs1131620 | 0.0002 | 0.90 | 0.21 (rec) | 0.02 |
| **THBS1** | rs2725797 | 0.0001 | 1.87 | 0.67 (add) | 2.02 |
| **CD40** | rs1883832 | 0.0002 | 2.00 | 0.97 (dom) | 0.64 |
| **ACTN3** | rs1815739 | 0.0001 | 1.62 | 1.11 (add) | 1.00 |
| **TCTEX1D1** | rs1060575 | 0.0004 | 2.39 | 1.24 (add) | 0.49 |

Of note are the differences between the current findings and our initial report of associations with *LTBP4* and *THBS1*, which was based on a subset (N=253) of the current cohort; 174 subjects overlap between the original and current cohorts. 54 of the original 253 subjects were dropped in the current cohort based on *DMD* mutation class (see above); an additional 25 were dropped based on other filtering criteria as described above. If we consider these 54 nonoverlapping subjects alone, we obtain MODs of 4.8, 3.9 for rs710160 (*LTBP4*), rs2725797 (*THBS1*) respectively, and variance-components (recessive) –log_10_(p-values) of 6.2, 4.6; thus these non-overlapping subjects contributed substantially to the original findings. Not surprisingly, detection of any given modifier across patient cohorts with substantial differences, including differences in *DMD* mutation status, remains challenging (see ^27^). This means that appreciably larger sample sizes will be needed to reliably replicate modifier effects in humans across potentially heterogeneous sets of subjects.

The results of our study, taken together with previous studies, are consistent with the idea that there may be *many* genes that can modify the DMD phenotype. In this case, simple sampling variability from one dataset to another could also make direct replication across studies difficult, as the effects of different genes become salient in different samples, against different genetic and possibly environmental backgrounds, and perhaps under different data analytic strategies ^27^. If this is the case, simply “pooling” all data together for ever larger sample sizes may only serve to reduce the impact of any one modifier gene to undetectable levels. Studies that examine the interaction between large-effect monogenic mutations and small-effect modifier variants, such as the one described here, are few. It is notable that the associations of common SNPs detected with this study were all non-coding variants, consistent with the majority of GWAS-associated variants linked to human traits ^38^.

Given the impact of cohort selection on the detection of modifiers, and that there are no existing additional data sets comparably large or larger than our current cohort for use in a standard replication design, we have employed an alternative to replication for the next stage of evaluation of the reasonableness of these new regions associated with LOA. As described in the companion paper to this one [Flanigan et al., medRxiv, 2021], we selected 6 loci with PPLD scores of ≥0.4 for further exploration, using multiple lines of evidence for plausibility for a biologic role for the identified SNPs in regulating genes or pathways potentially relevant to muscle biology. As described in that companion paper, we systematically applied a search for features indicative of functional regulatory variation, for evidence of associated gene regulation by eQTL analysis, and for evidence of direct physical interaction via Hi-C interactions; the results of those analyses and the biological plausibility of the candidates are described therein. The data we present here suggest that even in a relatively small data set— albeit the largest studied in DMD to date—our approach to the analysis of DMD phenotype and SNP association may be an effective strategy for prioritizing genes for experimental verification.

## Data Availability

All data produced in the present study are available upon reasonable request to the authors

## ACKNOWLEDGMENTS

This work was supported by the National Institutes of Health (NINDS NS085238) to KMF, RBW, and VJV. The Genotype-Tissue Expression (GTEx) Project was supported by the Common Fund of the Office of the Director of the National Institutes of Health, and by NCI, NHGRI, NHLBI, NIDA, NIMH, and NINDS. The data used for the analyses described in this manuscript were obtained from the GTEx Portal using the GTEx Analysis Release V8 (dbGaP Accession phs000424.v8.p2). The authors wish to acknowledge the many members of the United Dystrophinopathy Project consortium who participated in the enrollment of subjects in the original historical UDP database.

## AUTHOR CONTRIBUTIONS

R.B.W., V.J.V. and K.M.F conceived and designed the study. R.B.W., V.J.V., D.M.D., M.W., R.A., L.A., M.M-C., K.M.F., J.B., SC.S., and the UDP Investigators acquired and/or analyzed the data. R.B.W., V.J.V., P.T.M., and K.M.F. drafted the manuscript, which was reviewed and revised by all of the named authors.

## ETHICAL APPROVAL

The research involving human subjects has been been performed in accordance with the Declaration of Helsinki and was approved by the Nationwide Children’s Hospital Institutional Review Board (IRB) under approval 0502HSE046. Informed consent was obtained from all subjects.

## CONFLICTS OF INTEREST

Nothing to report.

## SUPPLEMENTAL FIGURES

**Figure S1.**
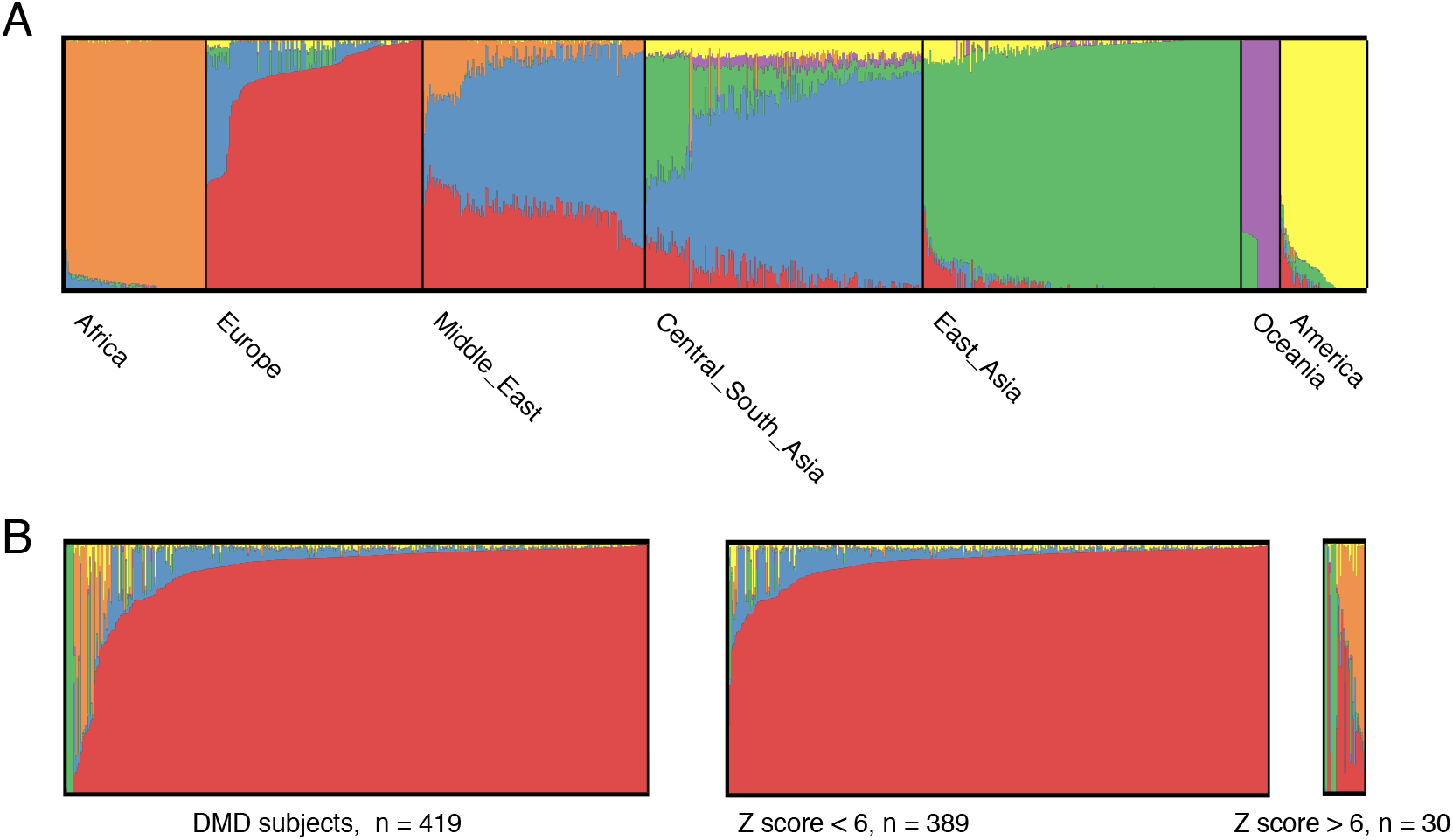
Z-score filtering for estimating ancestry of DMD subjects. **(A)** Plot of ADMIXTURE results for *K* = 6, 10/10 runs using the HGDP 938 reference panel. Each vertical line in the plot corresponds to one individual and colors represent 6 inferred ancestral populations. The color scheme roughly corresponds to geographic origins. **(B)** The 419 DMD study individuals were sub-divided by Z score threshold of 6 using the POPRES European ancestry reference space following Taliun *et al*.

**Figure S2.**
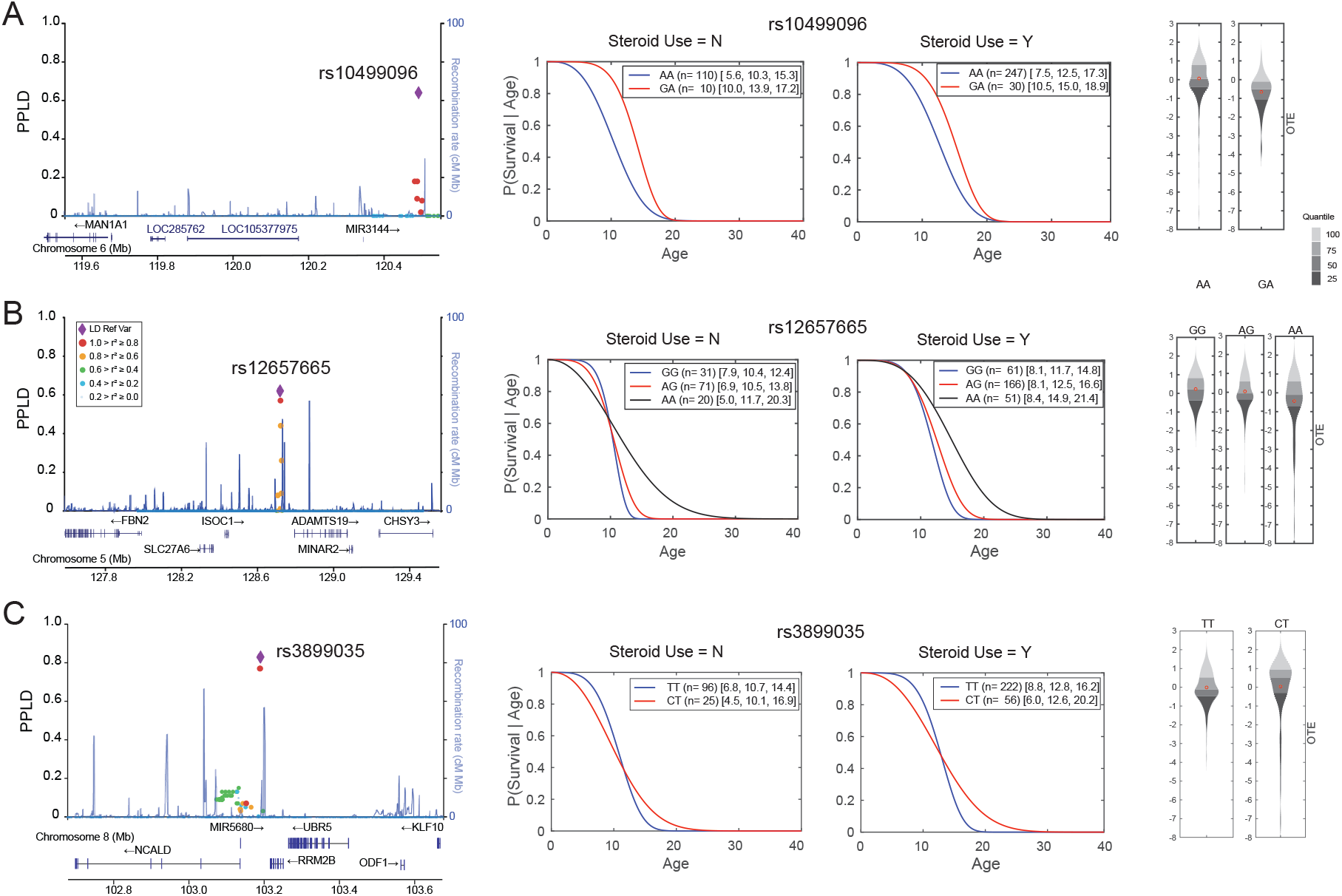
Additional regional plots of loci with PPLDs ≥ 40%. **(A)** through **(C)** are three additional regions from Table 1 with PPLDs ≥ 40%. The **left panel** shows regional LocusZoom plots of SNPs with their PPLD values shown on the primary y-axis and colored by their LD (*r*^*2*^, red ≥ 0.8, orange ≥ 0.6, green ≥ 0.4, light blue ≥ 0.2) with the lead SNP indicated with a purple diamond. The **middle panel** shows the estimated (Weibull) survival curves for the two Steroid (N/Y) groups, respectively; legends include the number n of individuals followed by [10^th^ percentile, median, 90^th^ percentile] of the age at LOA distribution. The **right panel** shows the density plots for the OTE residuals with the y-axis units in deviations in age at LOA from expectation, after adjusting for steroid use. Genotypes with <15 individuals are not shown, since our ability to accurately estimate the distribution in very small groups is limited.

## Supporting information

**Table S1. DMD exonic deletion/duplication mutations**.

**Table S2. DMD point mutations**.

**Table S3. SNP annotation of PPLD credible sets**.

## Notes

### Competing Interest Statement

The authors have declared no competing interest.

### Author Declarations

Ethics committee/IRB of Nationwide Children's Hospital and the University of Utah gave ethical approval for this work.

